# The importance of a consistent workflow to estimate muscle-tendon lengths based on joint angles from the conventional gait model

**DOI:** 10.1101/2021.03.10.21253257

**Authors:** Hans Kainz, Michael H Schwartz

## Abstract

**Background:** Musculoskeletal models enable us to estimate muscle-tendon length, which has been shown to improve clinical decision-making and outcomes in children with cerebral palsy. Most clinical gait analysis services, however, do not include muscle-tendon length estimation in their clinical routine. This is due, in part, to a lack of knowledge and trust in the musculoskeletal models, and to the complexity involved in the workflow to obtain the muscle-tendon length.

**Research question:** Can the joint angles obtained with the conventional gait model (CGM) be used to generate accurate muscle-tendon length estimates?

**Methods:** Three-dimensional motion capture data of 15 children with cerebral palsy and 15 typically developing children were retrospectively analyzed and used to estimate muscle-tendon length with the following four modelling frameworks: (1) 2392-OSM-IK-angles: standard OpenSim workflow including scaling, inverse kinematics and muscle analysis; (2) 2392-OSM-CGM-angle: generic 2392-OpenSim model driven with joint angles from the CGM; (3) modif-OSM-IK-angles: standard OpenSim workflow including inverse kinematics and a modified model with segment coordinate systems and joint degrees-of-freedom similar to the CGM; (4) modif-OSM-CGM-angles: modified model driven with joint angles from the CGM. Joint kinematics and muscle-tendon length were compared between the different modelling frameworks.

**Results:** Large differences in hip joint kinematics were observed between the CGM and the 2392-OpenSim model. The modif-OSM showed similar kinematics as the CGM. Muscle-tendon length obtained with modif-OSM-IK-angles and modif-OSM-CGM-angles were similar, whereas large differences in some muscle-tendon length were observed between 2392-OSM-IK-angles and 2392-OSM-CGM-angles.

**Significance:** The modif-OSM-CGM-angles framework enabled us to estimate muscle-tendon lengths without the need for scaling a musculoskeletal model and running inverse kinematics. Hence, muscle-tendon length estimates can be obtained simply, without the need for the complexity, knowledge and time required for musculoskeletal modeling and associated software. An instruction showing how the framework can be used in a clinical setting is provided on https://github.com/HansUniVie/MuscleLength.

## 1. Introduction

Three-dimensional gait analysis is used to objectively measure and identify gait abnormalities in people with movement disorders [1]. Most clinical gait laboratories use a variant of the Newington-Helen Hayes model [2,3], which was developed in 1980s and is now known as the conventional gait model (CGM). This model produces estimates of joint kinematics and kinetics. Data derived from gait analysis based on the CGM has been shown to change treatment decision-making in children with cerebral palsy [4].

Musculoskeletal models enable additional analyses beyond those obtained from the CGM. These additional analyses include the estimation of muscle-tendon length [5], muscle moment arms [6], muscle-tendon forces [7], and joint contact forces [8]. Although musculoskeletal models are as reliable as the CGM [9], and despite the above-mentioned advantages, most clinical gait laboratories do not routinely use them. Limited trust in the models, the required expertise, and extra time are likely the primary reasons for eschewing musculoskeletal modeling in many clinical gait laboratories.

Cerebral palsy (CP) is the most common paediatric neurologic disorder [10]. CP causes a progressive variety of musculoskeletal impairments, including bony torsions that alter muscle moment arms, and contractures that alter muscle-tendon lengths. Walking impairments due to the neuromusculoskeletal abnormalities are among the most profound disabilities in children with CP [11]. Orthopaedic surgerises, including muscle-tendon lengthening [12] or transfer [13], are used to improve walking in children with CP. Research has shown that pre-operative muscle-tendon length estimation based on musculoskeletal modelling can improve clinical decision-making in children with CP [5,14].

Two approoaches have been used to estimate muscle-tendon length based on musculoskeletal models. The standard musculoskeletal modelling approach (MSM approach) includes scaling a generic musculoskeletal model to the anthropometry of the individual, calculating joint angles via inverse kinematics, and extracting muscle-tendon lengths [15]. Alternatively, the joint angles obtained with the CGM can be used to drive a musculoskeletal model (CGM approach) and extract the muscle-tendon lengths [5,16,17]. The CGM approach has the advantage of skipping the scaling and inverse kinematic steps, which are needed for the MSM approach. However, the CGM approach potentially introduces inconsistencies between joint angles and muscle-tendon lengths due to the difference in segment reference frames and joint degrees-of-freedom between the CGM and most musculoskeletal models [18]. Furthermore, joint angles in the CGM are calculated with direct kinematics, whereas most musculoskeletal models use inverse kinematics for estimating joint angles, introducing another difference between the MSM and CGM approaches. So far, no studies evaluated the impact of the different approaches on estimated muscle-tendon lengths.

The aims of this study were to assess the impact of different approaches on estimated muscle-tendon lengths and provide the clinical and research community an simple method for estimating muscle-tendon length. We hypothesized that

H1 Muscle-tendon length will differ between the MSM and CGM approaches for some muscles due to different segment coordinate systems and joint degrees-of-freedom [18].
H2 Modifying a musculoskeletal model to have segment reference frames and joint degrees-of-freedom consistent with the CGM will lead to an improved agreement between the MSM and CGM approaches.
H3 Scaling and inverse kinematics is not necessary to get reasonable muscle-tendon length estimations if the joint angles from the CGM are used to drive an appropriate musculoskeletal model.

In the long run, we hope our research will motivate more clinical gait laboratories to include muscle-tendon length estimations in their clinical routine and, therefore, increase the number of positive treatment outcomes in children with CP.

## 2. Methods

### 2.1 Participants

Three-dimensional motion capture data of 15 children with bilateral CP (mean ± standard deviation height: 1.45±0.16 m, weight: 41.6±13.7 kg, metadata of one participant with CP did not include height and weight information) and 15 typically developing (TD) children (height: 1.37±0.22 m, weight: 37.7±9,1 kg) were retrospectively analyzed for this study. Data was collected at the Gillette Children’s Specialty Healthcare Center for Gait and Motion Analysis. The use of previously collected de-identified data was approved by the Institutional Review Boards. The motion capture data included marker trajectories of the Vicon Plug-in-Gait marker set for the lower limbs plus two additional markers on each thigh and one (TD participants) or two (participants with CP) additional markers at each shank.

### 2.2 Modelling frameworks

Four different modelling frameworks based on the CGM and two different musculoskeletal models were used to calculate muscle-tendon lengths (Fig. 1).

**Fig. 1.**
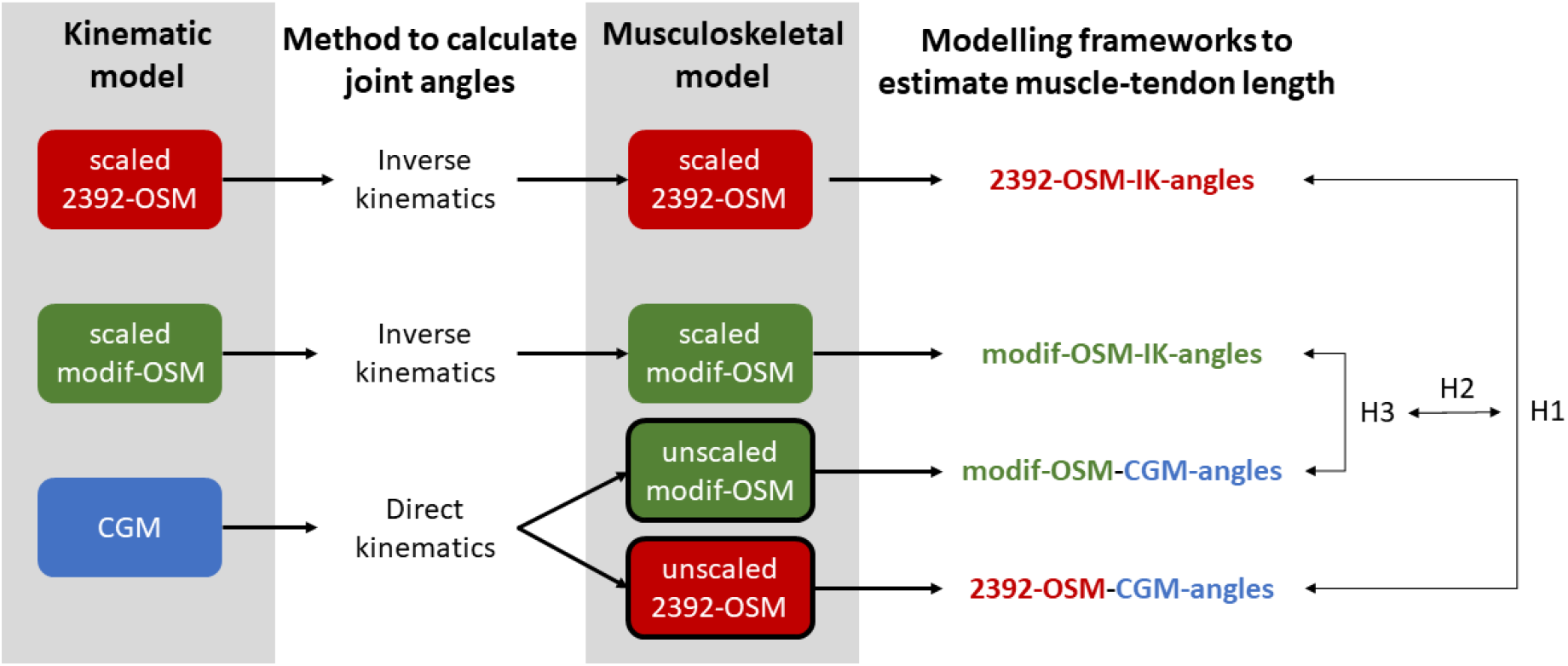
Modelling frameworks used to estimate muscle-tendon length. The CGM and two musculoskeletal OpenSim models were used to calculate joint kinematics. Afterward, these joint angles were used to drive scaled and unscaled musculoskeletal models to obtain muscle-tendon lengths. The right part of the figure indicates which modelling frameworks were compared to answer our research hypotheses. H1: muscle-tendon length for some muscles will be different between both previously used approaches to estimate muscle-tendon lengths. H2: modifying a musculoskeletal model will lead to an improved agreement between both approaches. H3: scaling and inverse kinematics is not necessary to get reasonable muscle-tendon length estimations if the joint angles from the CGM are used to run an appropriate musculoskeletal model.

#### 2.2.1 Joint kinematics calculations

Joint angles were calculated using the following three models:

##### CGM

The CGM [2,3] uses direct kinematics to calculate joint angles and is implemented in the Vicon Plug-in-Gait model, which is widely used in the clinical gait analysis community. This model outputs three rotations for the pelvis, hip and knee joint and one rotation (dorsi-/plantarflexion) for the ankle joint. Vicon Nexus was used for processing the data and calculating joint angles.

##### 2392-OSM

The 2392-OpenSim model [19], a widely used musculoskeletal model, was scaled to the anthropometry of each participant based on surface marker locations and estimated joint centres [20]. Afterwards, inverse kinematics was used to calculate joint angles. This model allows three rotational degrees-of-freedom at the hip joint and one degree-of-freedom at the knee joint. The subtalar and metatarsal joints were locked due to an insufficient number of foot markers and therefore only one degree-of-freedom (dorsi-/plantarflexion) was allowed at the ankle joint. The torso segment was removed from the model because we were only interested in muscle-tendon length of the lower extremities.

##### modif-OSM

The modified OpenSim model is based on the 2392-OSM, but included two additional degrees-of-freedom at each knee joint. Furthermore, the pelvic segment coordinate system was tilted by 13 degrees. These two modifications have been shown to improve the agreement in joint kinematic estimates between the CGM and musculoskeletal OpenSim models [9]. The same standard OpenSim workflow as described above, including scaling and inverse kinematics, was used to calculate joint angles.

#### 2.2.2 Muscle-tendon lengths calculations

Based on the joint kinematics obtained from the above mentioned three models, the following four modelling frameworks were used to calculate muscle-tendon lengths (Fig. 1, supplementary Table S1). In every framework, the analyze tool in OpenSim 4.1 [21] was used to derive muscle-tendon lengths.

##### 2392-OSM-IK-angles

framework uses the scaled 2392-OSM model to obtain joint angles and muscle-tendon lengths.

##### modif-OSM-IK-angles

framework uses the scaled modif-OSM model to obtain joint angles and muscle tendon lengths.

##### modif-OSM-CGM-angles

framework uses the unscaled modif-OSM model to calculate muscle tendon lengths. The model was driven by the joint angles obtained from the CGM.

##### 2392-OSM-CGM-angles

framework uses the unscaled 2392-OSM to calculate muscle tendon lengths. The model was driven by the joint angles obtained from the CGM.

For the modif-OSM-CGM-angles and 2392-OSM-CGM-angles frameworks, the joint angles from the CGM were exported from the .c3d file and a .mot file was created using a customized Matlab script, which is freely available on github (https://github.com/HansUniVie/MuscleLength). The .mot file is necessary to drive the OpenSim models.

### 2.3 Data analysis

Muscle-tendon lengths of the following muscles from the right leg were compared between the different modelling frameworks: semitendinosus, semimembranosus, biceps femoris (long head), rectus femoris, medial gastrocnemius, soleus, iliopsoas and adductor longus. These muscles were chosen because they are often included in soft tissue surgeries in children with CP. Data from children with CP and TD children were analyzed separately.

To address our first hypothesis, i.e. muscle-tendon length for some muscles will be different between both previously published approaches to estimate muscle-tendon length, we compared muscle-tendon length between the 2392-OSM-IK-angles and 2392-OSM-CGM-angles modelling framework using Statistical Parametric Mapping (SPM) [22]. To address our second hypothesis, i.e. modifying a musculoskeletal model will lead to an improved agreement between both approaches, we compared root-mean-square-differences (RMSD) and maximum differences (max-diff) between the 2392-OSM-IK-angles vs. 2392-OSM-CGM-angles frameworks to those between the modif-OSM-IK-angles vs. modif-OSM-CGM-angles frameworks. Paired t-tests were used to evaluate if there was a significantly improved agreement between modelling frameworks. For our third hypothesis, that scaling and inverse kinematics is not necessary to get reasonable muscle-tendon length estimations if the joint angles from the CGM are used to run an appropriate musculoskeletal model, we compared muscle-tendon length between the modif-OSM-IK-angles vs. modif-OSM-CGM-angles modelling frameworks using SPM. The Bonferroni correction for multiple comparisons was used to adjust the significance levels in the SPM analyses and paired t-tests.

Additionally, we visualized joint angles and quantified differences in joint angles between modelling frameworks using RMSD. All muscle-tendon lengths were normalized to the anatomical neutral position. Furthermore, joint angles and muscle-tendon lengths were time-normalized to 100 % of the gait cycle. To evaluate if different approaches have an impact on clinical reasoning, we quantified the number of CP children with short muscle-tendon length for each modelling frameworks. Short muscle-tendon length 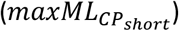 was defined as a maximum length during walking (*maxML*) below the mean 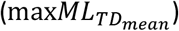 minus two standard deviation values 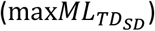 of the maximum length from the TD participants (Equation 1). Typical muscle-tendon length 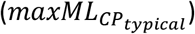 was defined as a maximum length equal to or above this threshold (Equation 2).

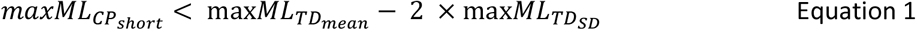

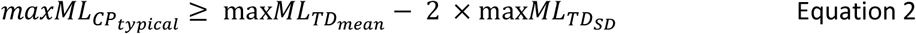

## 3. Results

### 3.1 Joint kinematics

Joint angles obtained with the 2392-OSM showed up to 17.1° (18.1°) RMSD (mean (standard deviation)) for hip flexion/extension angles compared to the other two models (CGM and modif-OSM). Knee and ankle joint kinematics in the sagittal plane were comparable between models with mean RMSD between 1.6° and 5.2° (Fig. 2, supplementary Table S2). Apart of hip and knee angles in the transverse plane, we found a good agreement in joint kinematics between the CGM and modif-OSM (mean RMSD between 0.5° and 7.1°, supplementary Table S2).

**Fig. 2.**
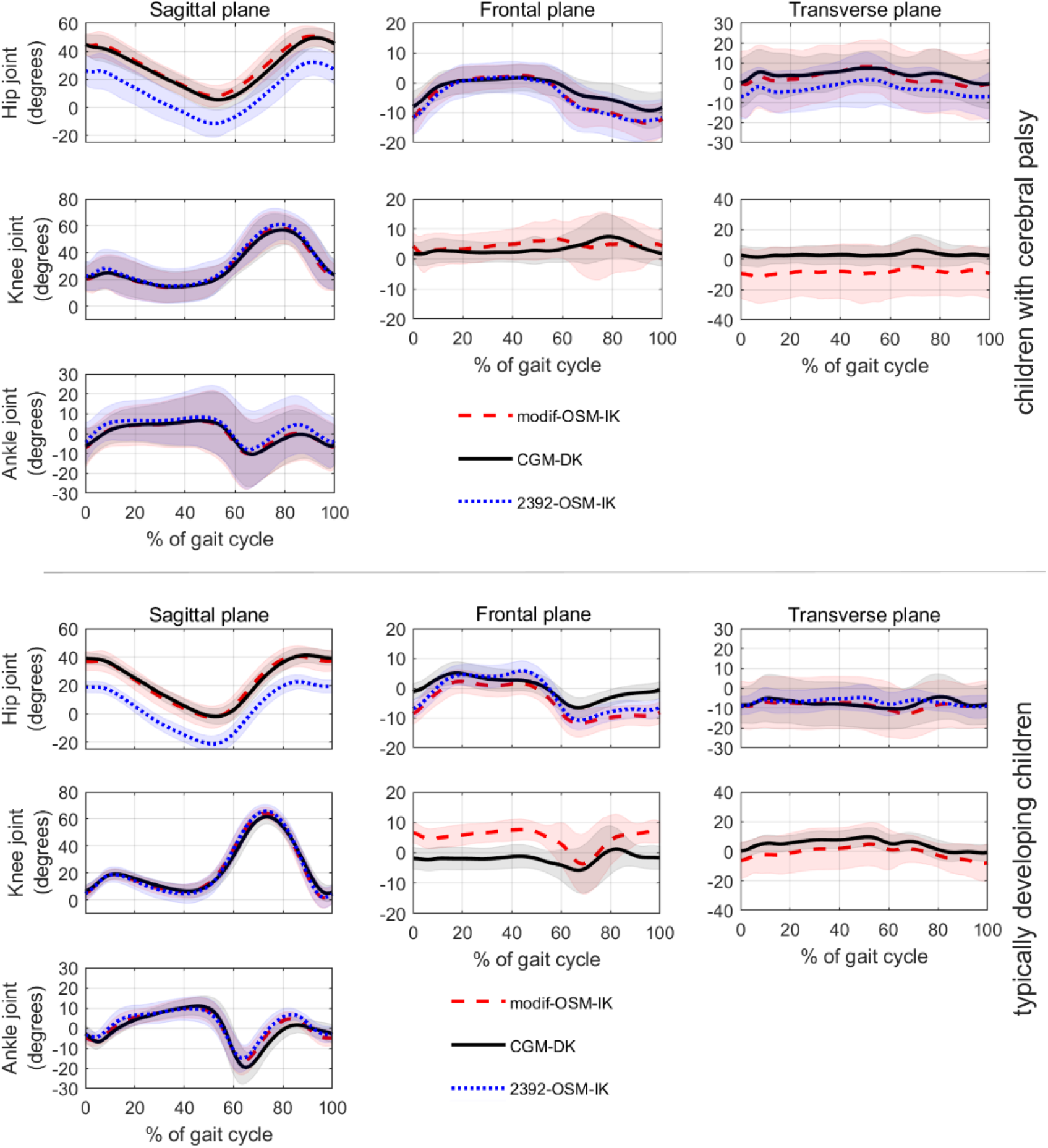
Joint kinematics (mean waveforms ± one standard deviation) obtained with the three different models for our participants with CP (top three rows) and our TD participants (button three rows). In both participants groups, hip flexion/extension angles from the 2392-OSM showed large differences compared to the other two models (CGM and modif-OSM).

### 3.2 Muscle-tendon lengths

In both participant groups (CP and TD), comparing muscle-tendon lengths between 2392-OSM-IK-angles and 2392-OSM-CGM-angles showed significant differences across the whole gait cycle for all analyzed muscles except the medial gastrocnemius and soleus muscles (Fig. 3). The comparison between modif-OSM-IK-angles and modif-OSM-CGM-angles only showed significant differences during short periods of the gait cycle.

**Fig. 3.**
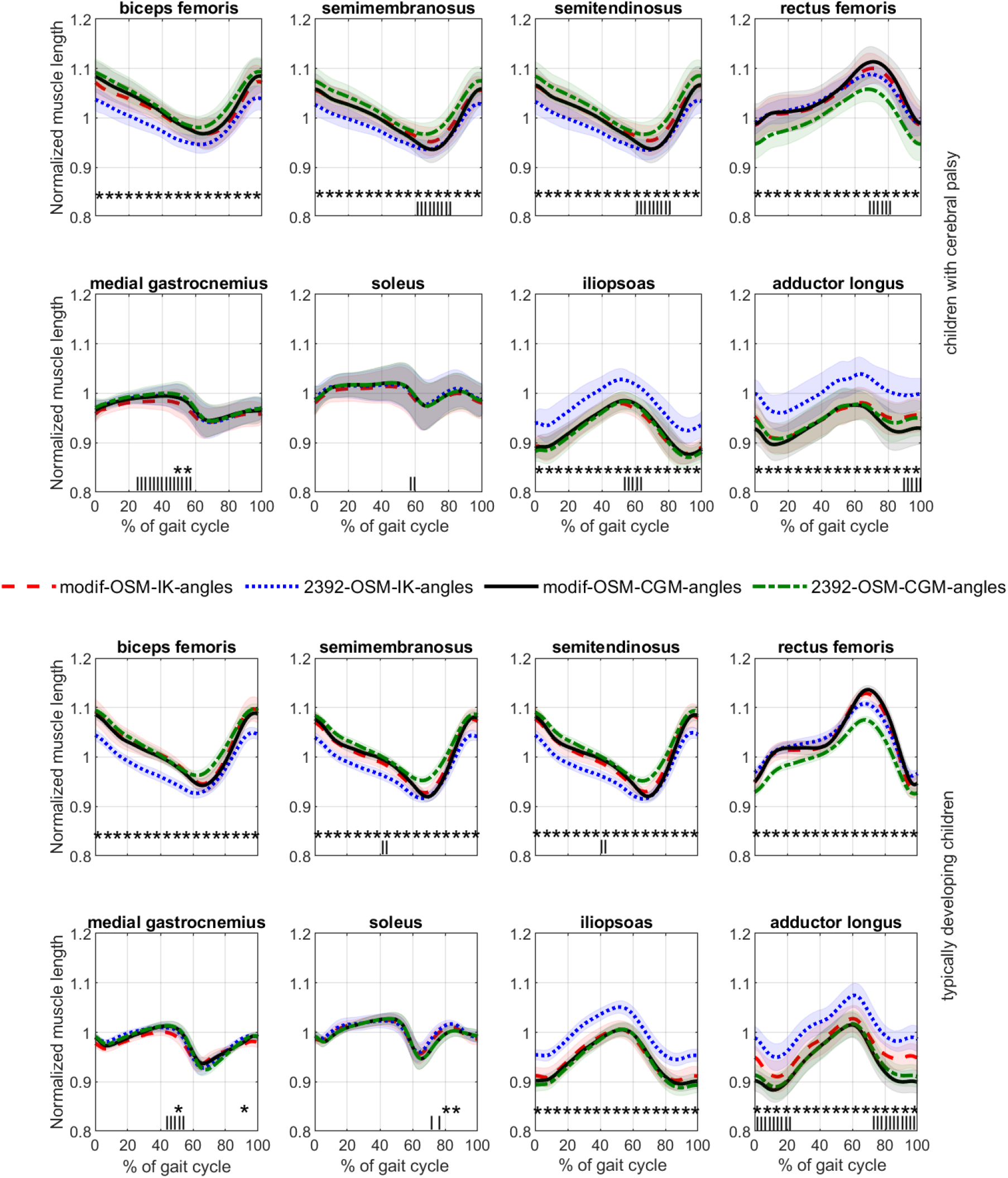
Normalized muscle length (mean waveforms ± one standard deviation) obtained with the four different modelling frameworks for our participants with CP (top two rows) and our TD participants (button two rows). *** = significant difference between 2392-OSM-IK-angles and 2392-OSM-CGM-angles, ||| = significant difference between modif-OSM-IK-angles and modif-OSM-CGM-angles.

In children with CP, average RMSD and max-diff in normalized muscle-tendon length for the 2392-OSM-IK-angles vs. 2392-OSM-CGM-angles comparison were 3.6% and 4.7%, respectively. Similar values were found in our TD participants (3.8% and 5.0% for average RMSD and max-diff, respectively). Average RMSD for the modif-OSM-IK-angles vs. modif-OSM-CGM-angles comparison were 1.3% in children with CP and 1.5% in TD participants. Mean max-diff were 2.3% and 2.7% in CP and TD participants, respectively. For most analyzed muscle-lengths, RMSD and max-diff were significantly lower for the modif-OSM-IK-angles vs. modif-OSM-CGM-angles comparison than for the 2392-OSM-IK-angles vs. 2392-OSM-CGM-angles comparison (Fig. 4).

**Fig. 4.**
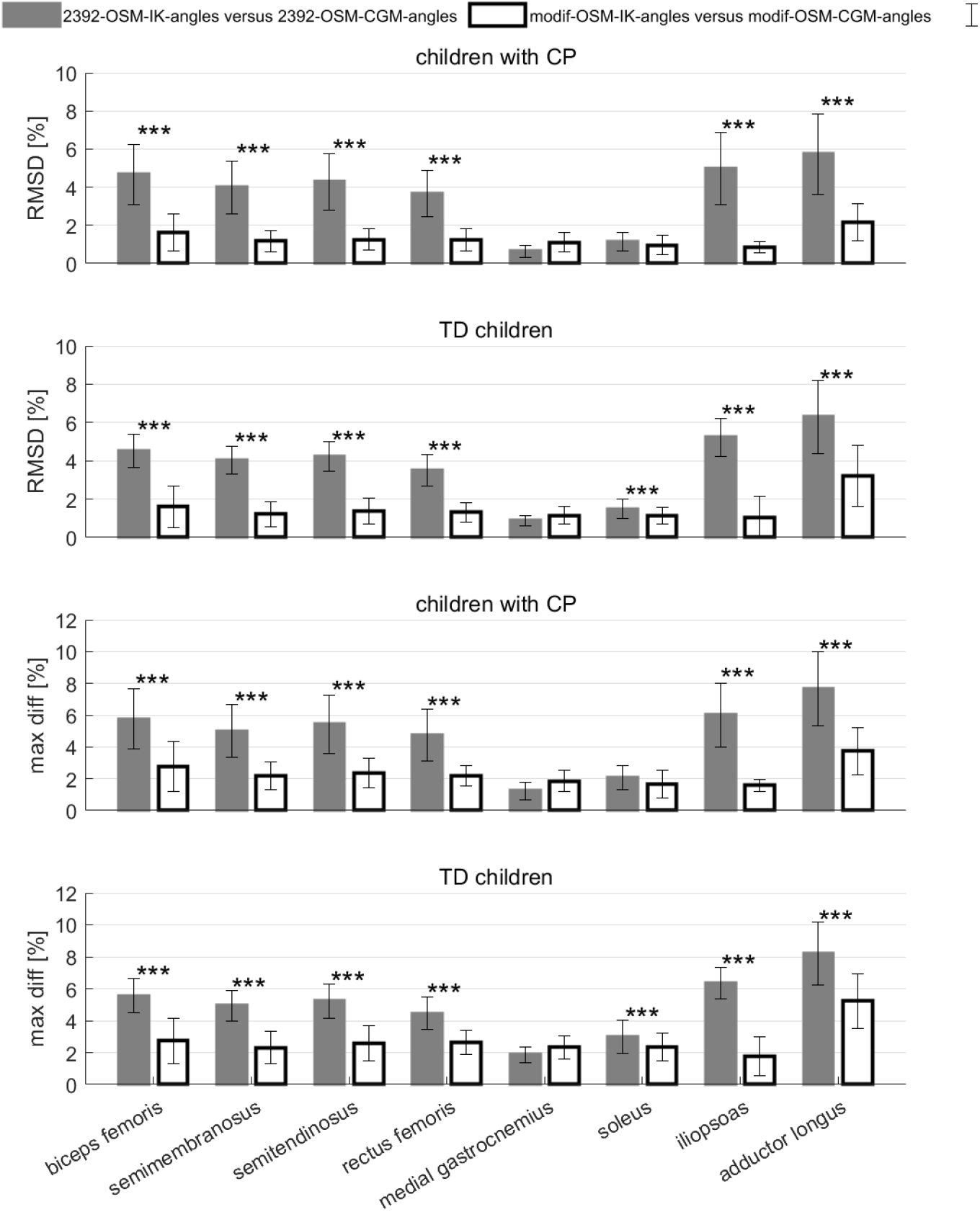
Root-mean-square-differences (RMSD) and maximum differences (max diff) in normalized muscle-tendon length between the 2392-OSM-IK-angles and 2392-OSM-CGM-angles (solid bards) and between the modif-OSM-IK-angles and modif-OSM-CGM-angles (empty bars). Error bars represent ± one standard deviation. RMSD and maximum differences decreased when using the modif-OSM compared to the 2392-OSM, which was in agreement with our second hypothesis. *** indicates significant differences (P<0.001).

### 3.3 Clinical reasoning

The difference in muscle-tendon lengths between participants with CP and the average TD values were similar between modelling approaches for many participants as long as a consistent approach was used for calculating muscle-tendon length of the CP and TD children (Fig. 5 and supplementary Fig. S1). Nevertheless, the number of children with short muscle-tendon length differed by up to 27% between modelling approaches (Fig. 6).

**Fig. 5.**
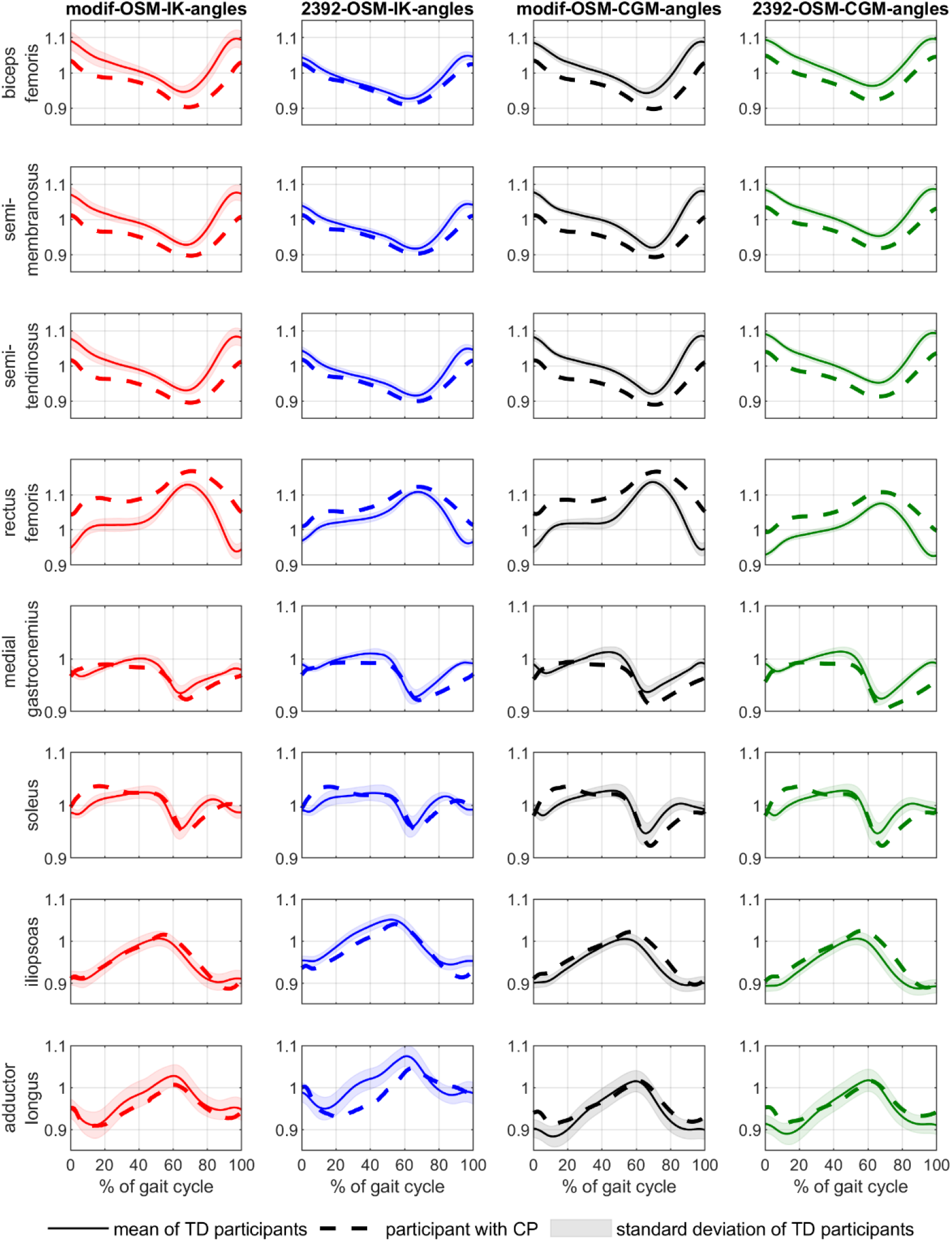
Comparison of muscle-tendon length from one of our participants with CP with the mean values of our TD participants using all four modelling approaches. Clinical interpretation based on the muscle-tendon length from the modif-OSM-IK-angles, modif-OSM-CGM-angles and 2392-OSM-CGM-angles modelling frameworks would be similar. The 2392-OSM-IK-angles approach showed compared to the other modelling frameworks smaller differences between the CP child and average TD waveforms for all analyzed muscles apart of the adductor longus muscle.

**Fig. 6.**
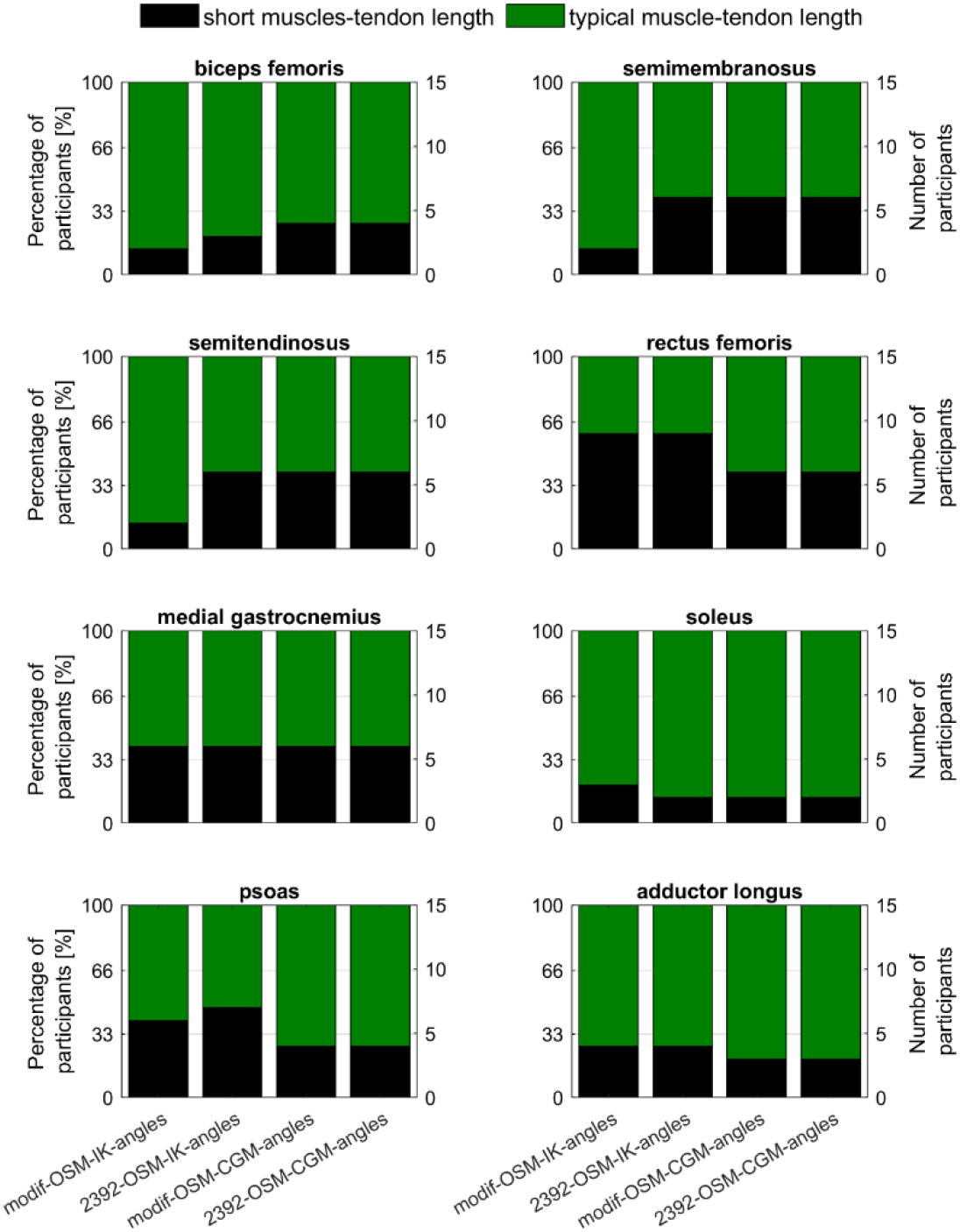
Percentage and number of participants with short and typical muscle-tendon length based on calculations with all four modelling frameworks.

## 4. Discussion

The aim of this study was to evaluate the impact of different modelling frameworks on the estimation of muscle-tendon lengths. In agreement with our first hypothesis, our findings showed that most muscle-tendon lengths exhibit large differences between the previously used and published approaches (2392-OSM-IK-angles vs. 2392-OSM-CGM-angles). Slightly modifying the musculoskeletal model led to a significant improvement in the agreement between the approaches and therefore confirmed our second hypothesis. Comparing the modif-OSM-IK-angles with the modif-OSM-CGM-angles framework showed similar muscle-tendon length waveforms with small RMSD and significant differences only during short periods of the gait cycle and, therefore, confirmed our third hypothesis.

Many clinical gait laboratories only use musculoskeletal modelling to estimate muscle-tendon lengths, and are not interested in other modeling capabilities, such as dynamic simulation or muscle force estimation. The first-author of this paper (HK) was involved in the organization of several international musculoskeletal OpenSim workshops. During these workshops many attendees from clinical gait laboratories were mainly interested in the estimation of muscle-tendon lengths for clinical decision-making in people with movement disorders. In addition to the limited trust, two technical barriers hindered most people to implement muscle-length estimations in their clinical routine. To the surprise of the organizers, the bottlenecks for many attendees were converting the .c3d files to OpenSim file formats (.trc and .mot files) and scaling the musculoskeletal model. This discovery is what led to the study presented here. We hope that our paper and findings will increase the trust in musculoskeletal simulations and facilitate muscle-tendon lengths estimations for an increasing number of clinical gait laboratories.

Scaling a musculoskeletal model introduces errors [20]. These errors influence joint kinematics and musculoskeletal calculations such as muscle-tendon length estimations. We showed that the joint angles from the CGM can be used to estimate muscle-tendon lengths without the need for scaling and inverse kinematics (modif-OSM-CGM-angles framework). Our proposed approach has two main benefits. First, it reduces the time and complexity of the processes required to estimate muscle-tendon lengths. Second, it can be done by a person with limited OpenSim experience without increasing uncertainties in the muscle-tendon length estimations.

Joint kinematics based on the CGM were comparable to previous studies with TD children [23]. and children with CP [24]. As expected, hip flexion/extension angles were significantly different between the CGM and 2392-OSM due to the different definition of the pelvis segment coordinate system [18]. Sagittal plane kinematics showed a good agreement between the CGM and modif-OSM but transverse plane kinematics differed between the approaches. This was likely due to the limited degree-of-freedom at the ankle joint in the modif-OSM and the global optimization approach used during the inverse kinematics analysis. Including an additional foot marker would unlock the subtalar joint in the modif-OSM, which would likely improve the agreement in joint kinematics between the modif-OSM and CGM.

Muscle-tendon lengths from muscles involved in hip movements showed the largest differences between modelling frameworks, whereas the medial gastrocnemius and soleus muscle-tendon lengths were similar between all approaches. Hence, muscle-tendon length from studies, which used different methods to estimate muscle-tendon lengths should be compared with caution, especially if the muscle of interest is involved in hip movements. In general, our muscle-tendon lengths were similar to previous studies that included children with CP [25,26].

Clinical reasoning based on comparing muscle-tendon lengths from our participants with CP with the mean values of our TD children was similar between modelling frameworks for most muscles and participants (73% to 100%, Fig. 5 and 6). Hence, clinical decisions are unlikely to be influenced by the choice of framework as long as the same framework is used for estimating muscle-tendon length in the CP and TD children. Nevertheless, we recommend using a modelling framework that ensures consistency between the joint angles and muscle-tendon length estimations. Furthermore, muscle-tendon lengths from a patient should never be compared to control data obtained with a different modelling approach because it could lead to wrong interpretation of the data (Fig. S1).

This study included some limitations. First, we only included four modelling frameworks in this study and all muscle-tendon length estimations were based on two musculoskeletal OpenSim models. We used OpenSim because it is widely used and is freely available compared to other commercially available musculoskeletal modelling software. Using different models [27] or software packages (e.g., AnyBody [28]) might lead to slightly different results. The main findings, however, are unlikely to change with different models and software packages and, therefore, we feel confident that our recommendations can be applied for any chosen modelling framework. Second, we did not have a gold standard to assess the accuracy of muscle-tendon lengths estimations with the different modelling frameworks. Hence, we only evaluated the consistency between joint kinematics and muscle-tendon lengths estimations between the different approaches. Using a consistent kinematics and musculoskeletal model is important to minimize uncertainties and errors associated with musculoskeletal modelling. Third, clinical reasoning is usually based on a comprehensive assessment that includes gait analysis data, medical history of the child, and physical examinations. In contrast, we only compared muscle-tendon lengths between CP and TD participants. A comprehensive assessment between our participants with CP and the TD control participants was, however, not our aim and therefore beyond the scope of this study.

In summary, this was the first study which highlighted common pitfalls in estimating muscle-tendon lengths. Furthermore, we showed how small modifications of a musculoskeletal model can lead to large improvements in the consistency between joint kinematics and muscle-tendon length estimations. The main findings of this study related to the practical implementation of a framework to estimate muscle-tendon lengths can be summarized as follows:

1. **Use consistent models**: The chosen kinematic (e.g. CGM) and musculoskeletal models (e.g. OpenSim model) should be based on matching segment reference frames and joint degrees of freedom.
2. **Angles from the CGM can be used to estimate muscle-tendon lengths**: This approach saves time, effort, and complexity in a clinical gait laboratory and eliminates potential scaling errors due to an inexperienced OpenSim user.
3. **Only compare patient muscle-tendon lengths to control data based on the same modelling framework**: Using different modelling frameworks for the patient and control data could lead to wrong interpretation and therefore disastrous clinical interventions.

A step-by-step manual on how to implement the modif-OSM-CGM-angles approach is provided on https://github.com/HansUniVie/MuscleLength. We hope our paper will help clinical gait laboratories to add muscle-length estimations to their clinical routine and improve clinical-decision making in children with CP.

## Data Availability

The data are not publicly available due to them containing information that could compromise research participant privacy or consent. Explicit consent to release data was not obtained from the patients, and data were collected up to 15 years ago. Thus, the vast majority of patients cannot be asked to provide their consent for release of their data. The data that support the findings of this study are available from the last author (MHS) upon reasonable request and subject to data sharing agreements.

## SUPPLEMENTARY MATERIAL

**Table S1.** Overview of differences between the modelling frameworks. DK=direct kinematics; IK=inverse kinematics.

| Modelling framework | Joint rotations<br>(degrees-of-freedom) |  |  | Scaling<br>Yes/No | Kinematic<br>calculations<br>DK/IK |
| --- | --- | --- | --- | --- | --- |
|  | Hip | Knee | Ankle |  |  |
| 2392-OSM-IK-angles | 3 | 1 | 1 | Yes | IK |
| modif-OSM-IK-angles | 3 | 3 | 1 | Yes | IK |
| modif-OSM-CGM-angles | 3 | 3 | 1 | No | DK |
| 2392-OSM-CGM-angles | 3 | 1 | 1 | No | DK |

**Table S2.** Mean (standard deviation) root-mean-square-differences in joint kinematics for the comparison between the CGM and the 2392-OSM and the comparison between the CGM and the modif-OSM. Sag=sagittal plane; front=frontal plane; trans=transverse plane; CP=participants with cerebral palsy; TD=typically developing participants

| comparison<br>group | CGM versus 2392-OSM |  | CGM versus modif-OSM |  |
| --- | --- | --- | --- | --- |
|  | CP | TD | CP | TD |
| pelvis-sag | 20.3 (10.8) | 11.2 (5.6) | 1.7 (3.2) | 0.5 (5.0) |
| pelvis-front | 8.7 (5.7) | 3.8 (1.8) | 2.1 (2.6) | 1.0 (1.2) |
| pelvis-trans | 1.6 (1.6) | 0.8 (0.7) | 1.6 (2.0) | 0.6 (0.5) |
| hip-sag | 17.1 (18.1) | 7.3 (4.6) | 3.5 (5.4) | 1.3 (5.5) |
| hip-front | 4.8 (5.1) | 2.0 (2.1) | 3.6 (5.6) | 1.7 (2.6) |
| hip-trans | 11.8 (8.6) | 8.8 (5.5) | 11.5 (9.2) | 5.0 (7.5) |
| knee-sag | 5.2 (5.2) | 2.3 (2.4) | 4.0 (5.1) | 1.9 (2.5) |
| knee-front |  |  | 7.1 (8.8) | 4.1 (4.0) |
| knee-trans |  |  | 14.8 (13.9) | 12.7 (8.1) |
| ankle-sag | 5.1 (6.0) | 3.3 (4.0) | 2.8 (4.6) | 1.6 (3.5) |

**Fig. S1.**
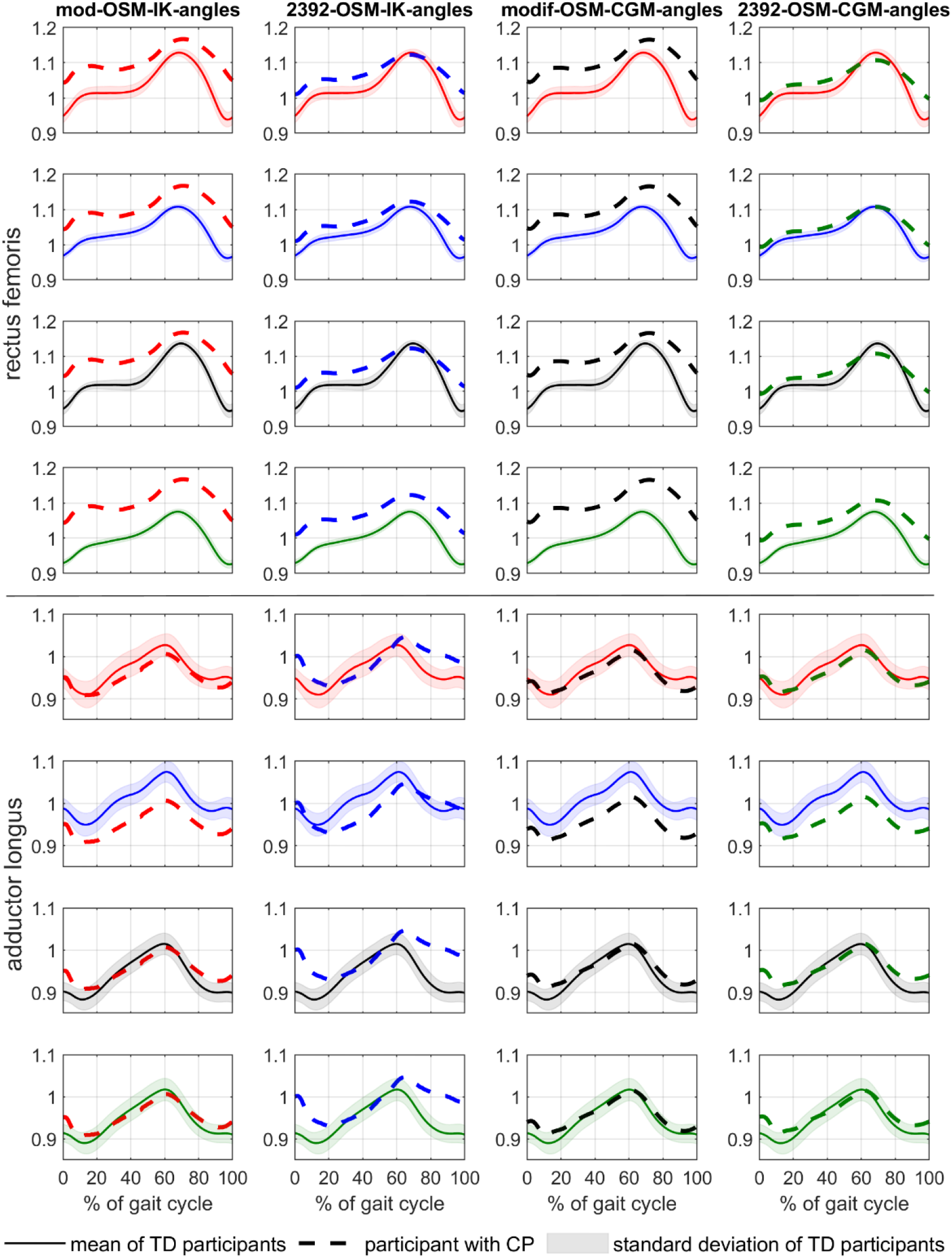
Comparison of rectus femoris (top four rows) and adductor longus (button four rows) muscle-tendon length from one of our participants with CP with the mean values of our TD participants using all four modelling approaches and different approaches for the reference values of the TD participants. Clinical interpretation would be affected based on the chosen reference data. E.g. first column row five and six → differences in adductor longus muscle-tendon length between the CP participant and TD values differed a lot. Data of the same participant with CP as in Fig. 5 are shown in this figure.

